# Cerebral microhemorrhages in children with congenital heart disease: Prevalence, risk factors, and impact on neurodevelopmental outcomes

**DOI:** 10.1101/2023.12.05.23299539

**Authors:** Kristen N. Andersen, Sicong Yao, Brian R. White, Marin Jacobwitz, Jake Breimann, Jharna Jahnavi, Alexander Schmidt, Wesley B. Baker, Tiffany S. Ko, J. William Gaynor, Arastoo Vossough, Rui Xiao, Daniel J. Licht, Evelyn K. Shih

**Affiliations:** Department of Pediatrics, Division of Neurology, Children’s Hospital of Philadelphia, Philadelphia, PA; Department of Pediatrics, Division of Biostatistics, The Children’s Hospital of Philadelphia, Philadelphia, PA 19104; Department of Pediatrics, Division of Cardiology, Children’s Hospital of Philadelphia and the Perelman School of Medicine at the University of Pennsylvania, Philadelphia, PA; Department of Anesthesiology and Critical Care Medicine, Children’s Hospital of Philadelphia, Philadelphia, PA 19104; Department of Surgery, Division of Cardiothoracic Surgery, The Children’s Hospital of Philadelphia, Philadelphia, PA; Department of Radiology, The Children’s Hospital of Philadelphia, Philadelphia, PA; Perinatal Pediatrics Institute, Children’s National Hospital, Washington D.C; Department of Neurology, University of Pennsylvania Perelman School of Medicine, Philadelphia, PA

## Abstract

**Background:** Infants with complex congenital heart disease (CHD) require life-saving corrective/palliative heart surgery in the first weeks of life. These infants are at risk for brain injury and poor neurodevelopmental outcomes. Cerebral microhemorrhages (CMH) are frequently seen after neonatal bypass heart surgery, but it remains unknown if CMH are a benign finding or constitute injury. Herein, we investigate the risk factors for developing CMH and their clinical significance.

**Methods:** 192 infants with CHD undergoing corrective cardiac surgery with cardiopulmonary bypass (CPB) at a single institution were prospectively evaluated with pre-(n = 183) and/or postoperative (n = 162) brain magnetic resonance imaging (MRI). CMH severity was scored based on total number of microhemorrhages. Antenatal, perioperative, and postoperative candidate risk factors for CMH and neurodevelopmental (ND) outcomes were analyzed. Eighteen-month neurodevelopmental outcomes were assessed using the Bayley-III Scales of Infants and Toddler Development in a subset of patients (n = 82). Linear regression was used to analyze associations between risk factors or ND outcomes and presence/number of CMH.

**Results:** The most common CHD subtypes were hypoplastic left heart syndrome (HLHS) (37%) and transposition of the great arteries (TGA) (33%). Forty-two infants (23%) had CMH present on MRI before surgery and 137 infants (85%) post-surgery. No parameters evaluated were significant risk factors for preoperative CMH. In multivariate analysis, cardiopulmonary bypass (CPB) duration (p < 0.0001), use of extracorporeal membrane oxygenation (ECMO) support (p < 0.0005), postoperative seizure(s) (p < 0.03), and lower birth weight (p < 0.03) were associated with new or worsened CMH postoperatively. Higher CMH number was associated with lower scores on motor (p < 0.03) testing at 18 months.

**Conclusion:** CMH is a common imaging finding in infants with CHD with increased prevalence and severity after CPB and adverse impact on neurodevelopmental outcomes starting at a young age. Longer duration of CPB and need for postoperative ECMO were the most significant risk factors for developing CMH. However, presence of CMH on preoperative scans indicates non-surgical risk factors that are yet to be identified. Neuroprotective strategies to mitigate risk factors for CMH may improve neurodevelopmental outcomes in this vulnerable population.

## Introduction

Infants with complex congenital heart disease (CHD) often undergo corrective surgery with cardiopulmonary bypass (CPB) within the first weeks of life. These infants are at increased risk for brain immaturity at birth, disordered brain development, acquired brain injury, and neurodevelopmental impairments spanning early development to adolescence and into adulthood.^1–6^ However, the underlying causes of this brain injury are not fully understood, and the impact on prognosis in the individual patient is variable. White matter injury (WMI) is the best characterized and most extensively studied type of injury in this patient population.^7–9^ Risk factors for WMI have been well-characterized.^7,10,11^ In contrast, although cerebral microhemorrhages (CMH) are the most common finding on perioperative brain magnetic resonance imaging (MRI), there is limited available data on risk factors for and clinical significance of CMH. In this study, we aimed to identify the prevalence of, risk factors for, and impact of CMH on neurodevelopmental outcomes in a cohort of infants with CHD who underwent surgery with CPB within the first two weeks of life.

CMH are rounded hypointense foci of <5 mm best visualized on susceptibility weighted imaging (SWI) sequences on MRI but can also sometimes be seen on gradient-echo (GRE).^12^ In other contexts, these foci are most common in perivascular locations throughout the parenchyma and are thought to represent deposits of hemosiderin that are associated with previous hemorrhagic events.^12–14^ Acute microhemorrhages, as seen postoperatively in our cohort, may represent other blood degradation products (e.g., extracellular deoxyhemoglobin, methemoglobin), all of which demonstrate susceptibility. Although CMH have been attributed to microscopic embolic material from the bypass pump, this would not account for their presence prior to surgery. CMH have been well-studied in adult populations and are frequently observed in healthy aged individuals, as well as patients with traumatic brain injury (TBI), dementia, cerebral amyloid angiopathy (CAA), or cerebral autosomal dominant arteriopathy with subcortical infarcts and leukoencephalopathy (CADASIL).^12^ In TBI, CMH are thought to arise from forces causing axonal sheer and compromise of the vasa nervorum with resulting microhemorrhage. Age has been shown to be an independent risk factor for developing CMH with a 20-40% higher incidence of CMH in those over 65 years.^15^ When located in the lobar regions of the brain, CMH are part of the diagnostic criteria for cerebral amyloid angiopathy.^16^ Hypertension is another independent risk factor for CMH, most commonly resulting in lesions that are distributed in deep and infratentorial brain regions.^15^ Importantly, the presence of CMH in these populations is a clinically significant finding that is linked with cognitive impairment, gait disturbances, and risk of intraventricular hemorrhage.^12,17,18^

In children, there are significantly fewer studies examining the prevalence and impact of CMH. Two prior studies have investigated CMH in children with CHD; however, these were limited by MRIs performed at one year of age (thus remote from the surgical insult) and small sample size. In this study, we address this gap in knowledge by analyzing a large single-center cohort of infants with CHD who underwent corrective surgery with CPB within the first weeks of life. Our prospective study benefited from the collection of both pre- and postoperative imaging as well as the collection of long-term neurodevelopmental outcomes. Our goals were to: (1) determine the prevalence of cerebral microhemorrhage in both the preoperative and postoperative time periods; (2) elucidate antenatal preoperative and operative risk factors associated with cerebral microhemorrhages; and (3) investigate the impact of cerebral microhemorrhage on neurodevelopmental outcomes in this population.

## Methods

### Study Design and Population

We performed a post hoc analysis of data prospectively collected from 2008-2019 as part of a longitudinal cohort study of term infants with CHD who underwent neonatal cardiac surgery with CPB at the Children’s Hospital of Philadelphia (CHOP). This study was approved by the Institutional Review Board (IRB 11-008191) and all participating families provided informed consent. Full term neonates (born at ≥ 36 weeks estimated gestational age) who required cardiac surgery with cardiopulmonary bypass within the first two weeks of life were eligible for inclusion. Infants who were small for gestational age (<2kg) or who experienced neonatal depression (5-minute APGAR <5 or cord pH <7), failed to undergo surgery, experienced preoperative cardiac arrest requiring chest compressions, significant intracerebral hemorrhage, or showed evidence of end-organ injury (AST/ALT above twice normal, creatinine >2 mg/dL, or heart failure) were excluded. Patients underwent at least one prospective brain MRI (pre- or post-operatively).

Patient characteristics, surgical, and medical history from the neonatal hospitalization were prospectively collected in a REDCap (Research Electronic Data Capture, Nashville, TN) research database hosted at CHOP. Captured variables included patient demographics (sex, race, ethnicity, gestational age), birth information (type of delivery, birth weight, head circumference, maternal pregnancy complications), cardiac diagnoses, genetic syndromes, surgical management, and details of pre-, intra-, and postoperative clinical care. For analysis purposes, cardiac diagnosis was divided into three subgroups: hypoplastic left heart syndrome (HLHS), transposition of the great arteries (TGA), or other, due to the small numbers of patients with diagnoses other than HLHS and TGA. Note was made of patients who received preoperative cardiac catheter interventions, which included balloon atrial septostomy, atrial septal stent placement, and diagnostic catheter angiograms. Subgroups for race included Black, Asian, Caucasian, Native American/Pacific Islander, Mixed, other, or unknown. Ethnicity was collected as either Hispanic/Latino or other.

### Cardiac Surgery

All infants underwent corrective or palliative surgery with cardiopulmonary bypass (CPB) within the first two weeks of life. Surgery was performed with deep hypothermic circulatory arrest (DHCA) for all subjects requiring aortic arch repair. Normothermic continuous bypass strategies were used for all biventricular repairs. All patients received modified ultrafiltration (MUF) on termination of CPB as standard of care. The total duration of CPB, deep hypothermic cardiac arrest (DHCA), and cross-clamp were calculated from anesthesia and operative records. For patients receiving DHCA, the total CPB time was defined as the time from initiation of bypass until the start of DHCA plus the time from the end of DHCA to removal from CPB.

### Neuroimaging

Brain MRIs were performed under general endotracheal anesthesia immediately before the neonatal cardiac surgery and approximately one-week post-surgery. MRIs were performed on a single 1.5T Avanto MRI system (Siemens Medical Systems, Malvern, PA) with a 12-channel head coil. Brain MRI sequences included T1-weighted magnetization-prepared rapid acquisition gradient echo (T1-MPRAGE TR/TE/inversion time=1980/2.65/1100 milliseconds, flip angle=15°, voxel size=0.4×0.4×1.5 mm, matrix=256×256), T2-weighted sampling perfection with application-optimized contrasts using different flip angle evolution (T2-SPACE, TR/TE=3200/453 milliseconds, voxel size = 0.9×0.9×2 mm, matrix=256×254), diffusion-weighted imaging (DTI; TR/TE=2903/86 milliseconds; slice thickness=4 mm; b values=0, 1000mm/s2; 20 directions; matrix 128×128), and susceptibility-weighted imaging (SWI; TR/TE=49/40 milliseconds, slice thickness=2 mm, matrix=256×177). T1- and T2-weighted sequences were acquired in the axial plane and reformatted in the sagittal and coronal planes.

MRIs were reviewed by a pediatric neurologist (D.J.L.) with expertise in brain image interpretation. WMI was identified by T1 hyperintensity and was rated using the validated quartered point system (QPS) with increasing scale indicating increasing severity in WMI: 0 signifies no WMI, 1 mild WMI, 2 to 3 moderate WMI, and 4 severe WMI.^19^ Volumetric measurements of WMI were performed by manual segmentation using ITK/SNAP.^20^ Cerebral microhemorrhage severity was determined by the number of lesions counted on SWI. We considered cerebral microhemorrhage as both a dichotomous variable (either present or absent) and as a continuous variable (number of lesions). We considered both the number of cerebral microhemorrhages present at any given time point as well as the difference in cerebral microhemorrhage number between preoperative and postoperative MRIs. Preoperative MRIs were additionally evaluated for total maturation score (TMS), a previously validated semiquantitative scoring system used to assess whole brain maturity across four parameters: myelination, cortical infolding, involution of glial cell migration bands, and presence of germinal matrix tissue.^7^

### Neurodevelopmental Assessment

A subgroup of 82 patients with postoperative MRI returned for detailed neurodevelopmental assessment at 18 months of age, as part of their standard clinical care. Motor, language, and cognitive functions were evaluated using the Bayley-III Scales of Infants and Toddler Development, which were performed by a certified child psychologist. Weight, length, and head circumference were also measured on the day of testing. The composite scores for each domain of this test are calculated based on comparison to normative age-matched samples. The standardized Bayley’s mean score of 100 (SD 15) indicates mid-average function, <85 (1 SD below mean) indicates mild impairment and risk of developmental delay, and <70 (2 SD below the mean) indicates moderate to severe impairment.^21^

### Statistical Analysis

Summary statistics are reported as mean and standard deviation (SD) or medians and interquartile ranges (IQRs) for continuous variables, as appropriate, and counts and percentages for categorical variables. For each outcome, we first performed univariate analysis on each risk factor, and then considered the variables with p-value <0.2 for subsequent multivariate modeling using the stepwise selection approach to determine the final models. Two-sided p-values of less than 0.05 were considered statistically significant. We performed all statistical analyses using R (R Core Team, Vienna, Austria).

Our first analysis was to assess for preoperative risk factors that predicted the presence of cerebral microhemorrhage on preoperative MRI. Due to the low incidence of preoperative cerebral microhemorrhages, associations with lesion number were not examined. The association for each risk factor with presence of preoperative cerebral microhemorrhages was assessed using Fisher’s exact test for categorical variables or Wilcoxon rank sum test for continuous variables. We did not pursue multivariate modeling for preoperative cerebral microhemorrhage as none of the risk factors reached a p-value of 0.2 in the univariate analysis.

Next, we performed linear regression to identify risk factors that predicted the change in number of cerebral microhemorrhage from pre-to postoperative MRI with univariate and subsequent multivariate analyses. In the multivariate analysis, we included risk factors that reached a p-value of 0.2 in the univariate analysis, and also forced birth weight into the model as we thought it was an important clinical parameter due to prior evidence of predictive utility.^1^

Finally, we assessed the impact of postoperative cerebral microhemorrhages on neurodevelopmental outcomes measured at 18 months. We calculated the Spearman correlation coefficient between each neurodevelopmental outcome (composite scores on the cognition, language, and motor tests of the Bayley-III Scales of Infant and Toddler Development; weight, length, and head circumference) with postoperative cerebral microhemorrhage and additionally performed linear regression to adjust for covariates of interest, including sex, gestational age, duration of bypass, use of postoperative ECMO, and seizure. The impact of white matter injury (total WMI volume, QPS score, TMS) on neurodevelopment was assessed using simple linear regression.

## Results

### Patient characteristics

Of a total of 192 consented patients enrolled in the study, 183 (95.3%) received a preoperative brain MRI. Reasons for not receiving an MRI included: parents rescinded consent (n=2), medical instability (n=1), and unavailability of the MRI on the morning of surgery (n=6). One hundred and sixty-two patients had both a pre- and postoperative MRI, 21 had only a preoperative MRI, and 4 had only a postoperative MRI (Table 1, Figure 1). The median time from surgery to postoperative MRI was 7 days [IQR= 5-8]. A subset of patients with postoperative imaging (n=82) returned for neurodevelopmental assessment at 18 months of age and were evaluated using the Bayley-III Scales of Infants and Toddler Development.

**Table 1.** Baseline characteristics of infants with congenital heart disease (CHD) who underwent surgery with cardiopulmonary bypass (CPB), according to analysis group. MRI indicates magnetic resonance imaging; SWI, susceptibility weighted imaging; ND, neurodevelopmental outcomes; CHD, congenital heart disease; HLHS, hypoplastic left heart syndrome; TGA, transposition of the great arteries; WMI, white matter injury; QPS, quartered point system; TMS, total maturation score; CPB, cardiopulmonary bypass; DHCA, deep hypothermic circulatory arrest; ACT, activated clotting time; preop, preoperative; post, postoperative.

| **Patient Characteristics,**<br>**median [IQR] or n (%)** | **Preop MRI** | **Preop to Postop**<br>**MRI Comparison** | **ND outcomes** |
| --- | --- | --- | --- |
| **Demographics** | **N = 183** | **N = 162** | **N = 82** |
| Gestational age at birth, week | 39 [38-40] | 39 [38-39] | 39 [38-39] |
| Birth weight, kg | 3 [3-4] | 3 [3-4] | 3 [3-4] |
| Head Circumference, cm | 34 [33-35] | 34 (33-35) | 34 (33-35) |
| Female | 77 (42) | 68 (42) | 41 (50) |
| Caucasian | 138 (75) | 121 (75) | 62 (76) |
| Black | 21 (11) | 18 (11) | 9 (11) |
| Asian | 2 (1) | 4 (2) | 0 (0) |
| Latino | 13 (7) | 10 (6) | 5 (6) |
| Vaginal delivery | 106 (58) | 95 (59) | 52 (63) |
| **Cardiac Diagnosis** |  |  |  |
| HLHS | 68 (37) | 57 (35) | 27 (33) |
| TGA | 59 (32) | 53 (32) | 22 (27) |
| Other | 56 (31) | 51 (31) | 33 (40) |
| **Preoperative Factors** |  |  |  |
| Cardiac catheter intervention | 39 (21) | 35 (22) | 15 (18) |
| Arrhythmia | 12 (6.6) | 9 (5.6) | 6 (7) |
| Hemoglobin* (g/dL) | 15 [13-16] | 15 [13-16] | 14 [13-16] |
| Platelets** (10<sup>3</sup>/uL) | 268 [222-315] | 274 [226-318] | 294 [231-360] |
| Patients with WMI | 40 (22) | - | - |
| WMI volume | 0 [0-0] | - | - |
| QPS score | 0 [0-0] | - | - |
| TMS score | 10 [9-11] | - | - |
| **Surgical factors** |  |  |  |
| Age at surgery, days | 4 [3-5] | 4 [3-5] | 4 [3-6] |
| CPB duration, min | 49 [39-76] | 49 [39-74] | 45 [39-68] |
| Cross clamp duration, min | 45 [38-60] | 44 [37-59] | 42 [36-51] |
| DHCA duration, min | 33 [0-44] | 32 [0-43] | 33 [0-42] |
| Maximum ACT, sec | 861 [662-1500] | 887 [661-1500] | 805 [677-1318] |
| Intra-operative arrhythmia | 27 (15) | 20 (12) | 11 (13) |
| **Postoperative Factors** |  |  |  |
| ECMO support | 7 (3.8) | 4 (2.5) | 4 (5) |
| Seizure | 17 (9.3) | 16 (9.9) | 12 (15) |
| Cardiac arrest | 13 (7.1) | 10 (6.2) | 7 (9) |
| Hemoglobin* (g/dL) | 13 [11-15] | 13 [11-15] | 14 [11-16] |
| Platelets** (10<sup>3</sup>/uL) | 102 [75-163] | 102 [75-163] | 106 [78-157] |
| Patients with WMI | - | 37 (23) | 23 (28) |
| WMI volume | - | 19 [0-89] | 21 [0-124] |
| QPS score | - | 1 [0-2] | 1 [0-2] |

\*Hemoglobin values were collected from blood gas records within 72 hours of surgery
\*\*Platelet values were collected from blood gas records within 72 hours of surgery

**Figure 1.**
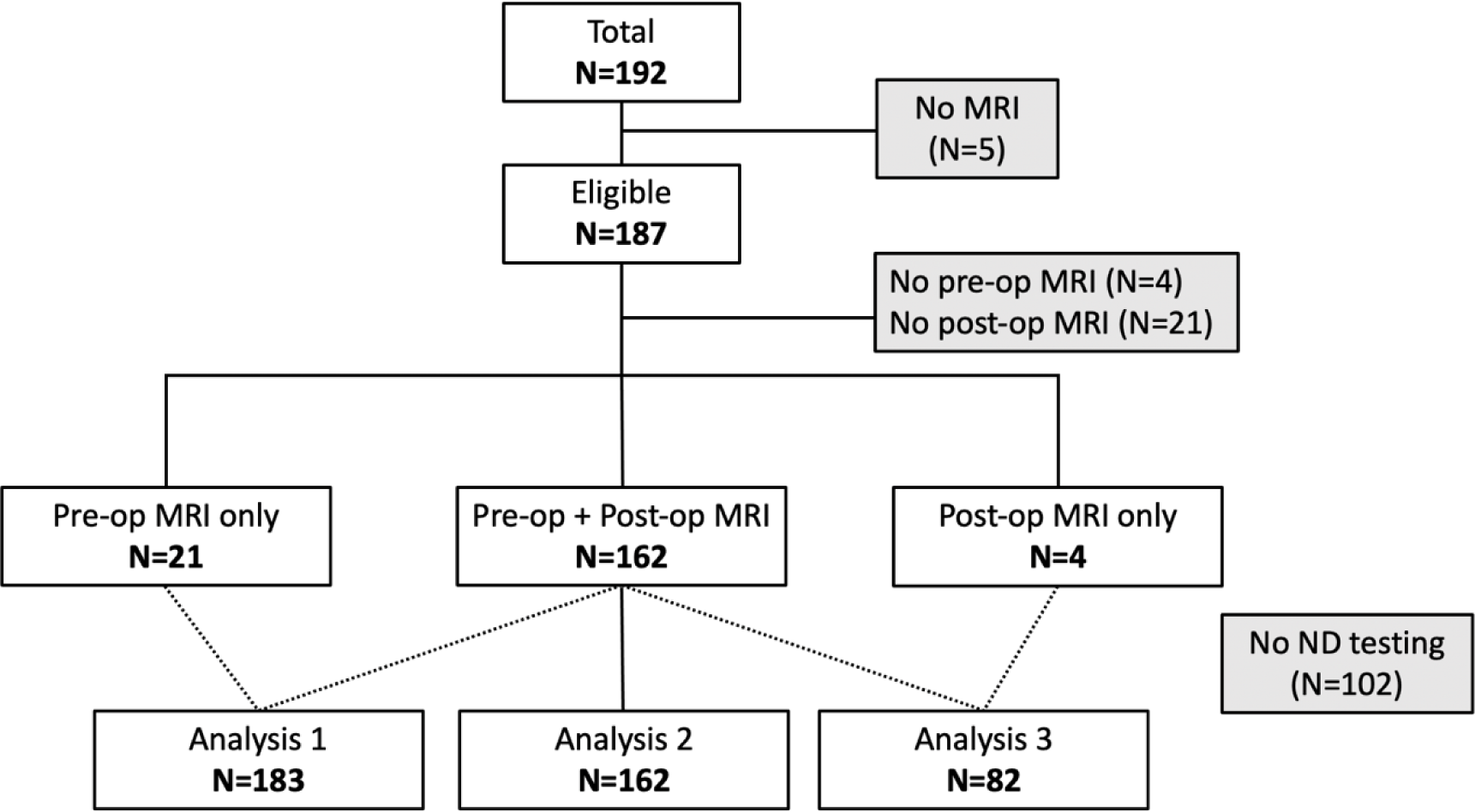
Flowchart of included patients in study analysis. Analysis 1 includes patients with preoperative MRI: Analysis 2, patients with pre- and postoperative MRI; Analysis 3, patients with postoperative MRI and neurodevelopmental outcomes. MRI indicates magnetic resonance imaging; ND, neurodevelopmental outcomes; pre-op, preoperative; post-op, postoperative.

The median age of patients at the time of cardiac surgery was 4 days [IQR = 3-5 days]. The majority of patients were Caucasian (n = 142; 76%) and a minority identified as Hispanic/Latino (n = 13; 7%). Sixty-nine infants (37%) had a diagnosis of hypoplastic left heart syndrome (HLHS) and 62 (33%) had transposition of the great arteries (TGA). The remaining 56 (30%) had structural heart defects that included aortic arch anomalies (total n=25; interrupted aortic arch n=8; aortic arch hypoplasia/coarctation n=8; aortic arch hypoplasia/coarctation with ventricular septal defect n=9), unbalanced common atrioventricular canal (n=7), tetralogy of Fallot (n=9), Ebstein’s anomaly (n=1), tricuspid atresia (n=1), right ventricular aorta with pulmonary atresia (n=1), truncus arteriosus (n=2), double inlet left ventricle (n=3), and double outlet right ventricle (n=7).

### Cerebral Microhemorrhage Prevalence and Distribution

Of 183 infants analyzed, 42 (23%) had one or more cerebral microhemorrhage present on preoperative MRI. The number of cerebral microhemorrhage in the preoperative subgroup ranged from 0-9 with a median of zero (Figure 2a). CMH were distributed throughout the brain in the posterior fossa (27%), deep gray matter structures (25%), and cerebral cortex/subcortical white matter junction (16%) (Figure 2b).

**Figure 2.**
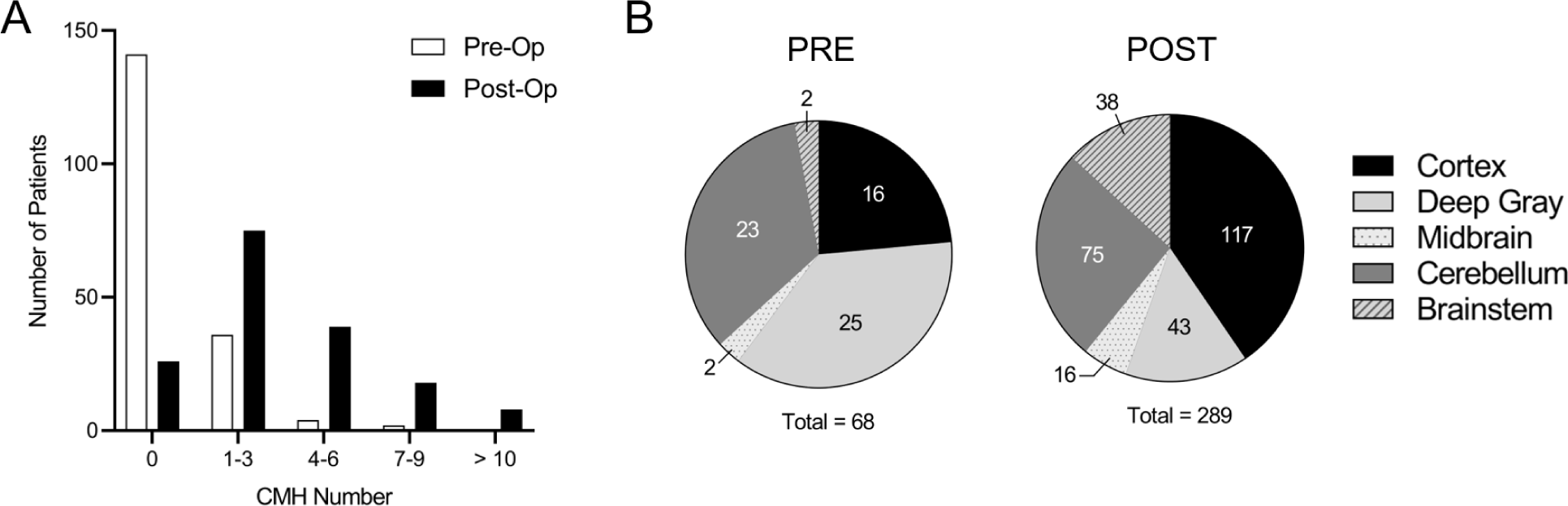
(a) Increased prevalence of cerebral microhemorrhage (CMH) on postoperative scans compared to preoperative imaging. White bars indicate patients with pre-operative imaging. Black bars indicate patients with postoperative imaging. **(b) Location of cerebral microhemorrhage (CMH) on preoperative vs postoperative brain magnetic resonance imaging (MRI).** Cortex refers to CMH in both the cortex proper and subcortical white matter junction. Pre indicates preoperative; Post, postoperative.

Of 162 infants with both pre- and postoperative scans, 137 (85%) had one or more CMH on postoperative MRI (Figure 2a). Of the patients in this group, the median number of CMH on postoperative MRI was 3 [IQR= 1-5] and the mean change in cerebral microhemorrhage number between pre- and postoperative MRI was 2 [IQR= 0-4] (representative MRIs shown in Figure 3). Postoperative CMH were located in the cerebral cortex/subcortical white matter junction (40%), posterior fossa (45%), and deep gray matter structures (15%) (Figure 2b).

**Figure 3.**
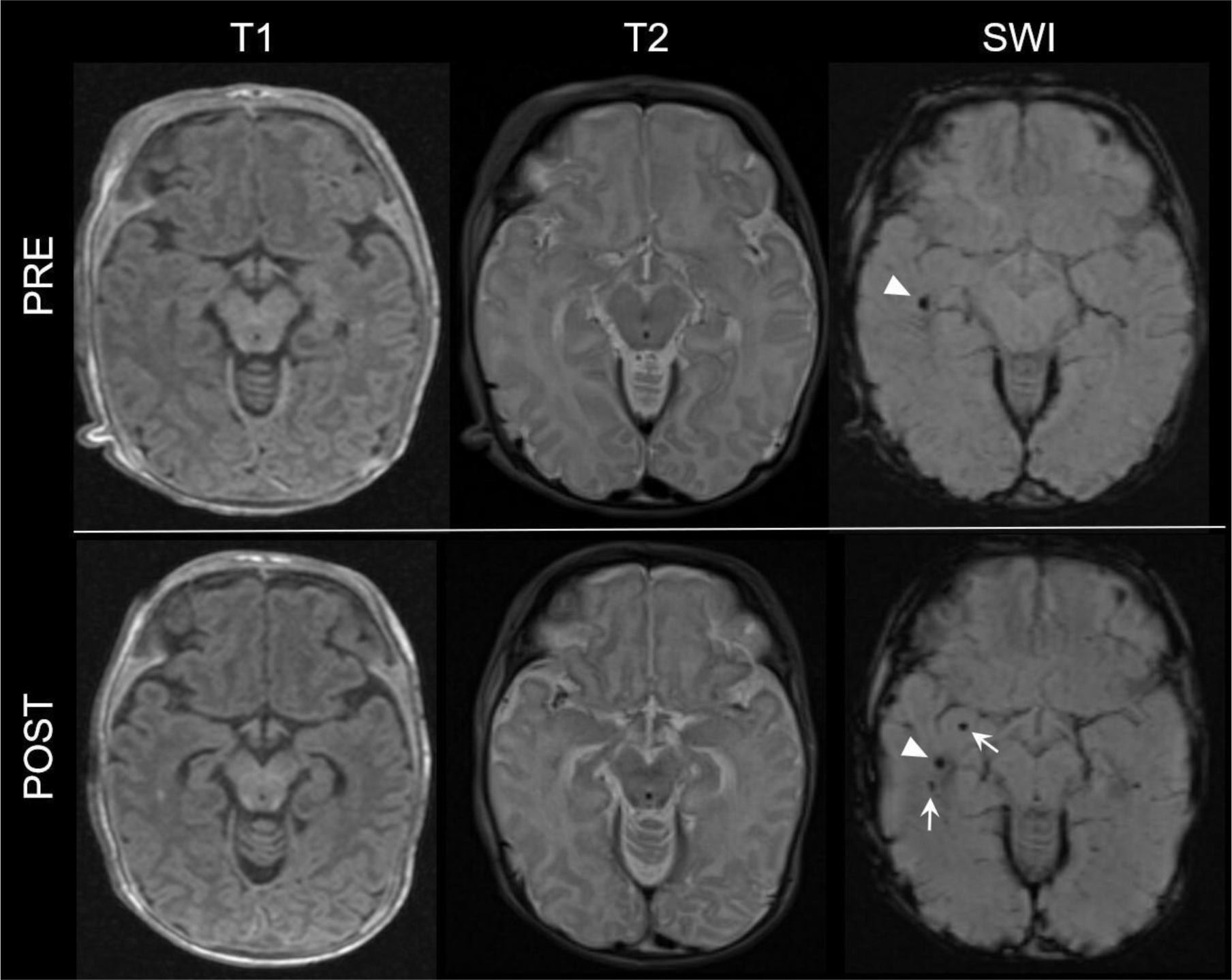
Representative magnetic resonance imaging (MRI) of a single patient with pre- and postoperative cerebral microhemorrhage. CMH are often not visible on T1-weighted or T2-weighted MRI but are visible on susceptibility weighted imaging (SWI). Arrowheads demarcate cerebral microhemorrhage present on both preoperative and postoperative SWI. Arrows highlight the accrual of additional CMH on postoperative SWI. T1 indicates T1-weighted MRI; T2, T2-weighted MRI; SWI, susceptibility weighted imaging; Pre, preoperative; Post, postoperative

### Risk Factors for Preoperative Microhemorrhages

The prevalence and severity of preoperative cerebral microhemorrhage was not affected by sex, gestational age, delivery type (vaginal vs. cesarean section), or cardiac diagnosis (Table 2). Additionally, there was no significant association with race, ethnicity, gestational factors (i.e., maternal hypertension, diabetes, preeclampsia), total maturation score (TMS), or preoperative arrhythmia. However, the small number of patients within each of these subgroups limited the ability to detect any potentially significant effects. Of 40 patients who had catheter interventions before surgery, 12 (30%) had cerebral microhemorrhage present, which was not statistically different from patients without such interventions (30/143 or 21% with cerebral microhemorrhage, p=0.29). Stroke as a consequence of balloon atrial septostomy (BAS) was not seen in this cohort.

**Table 2.**
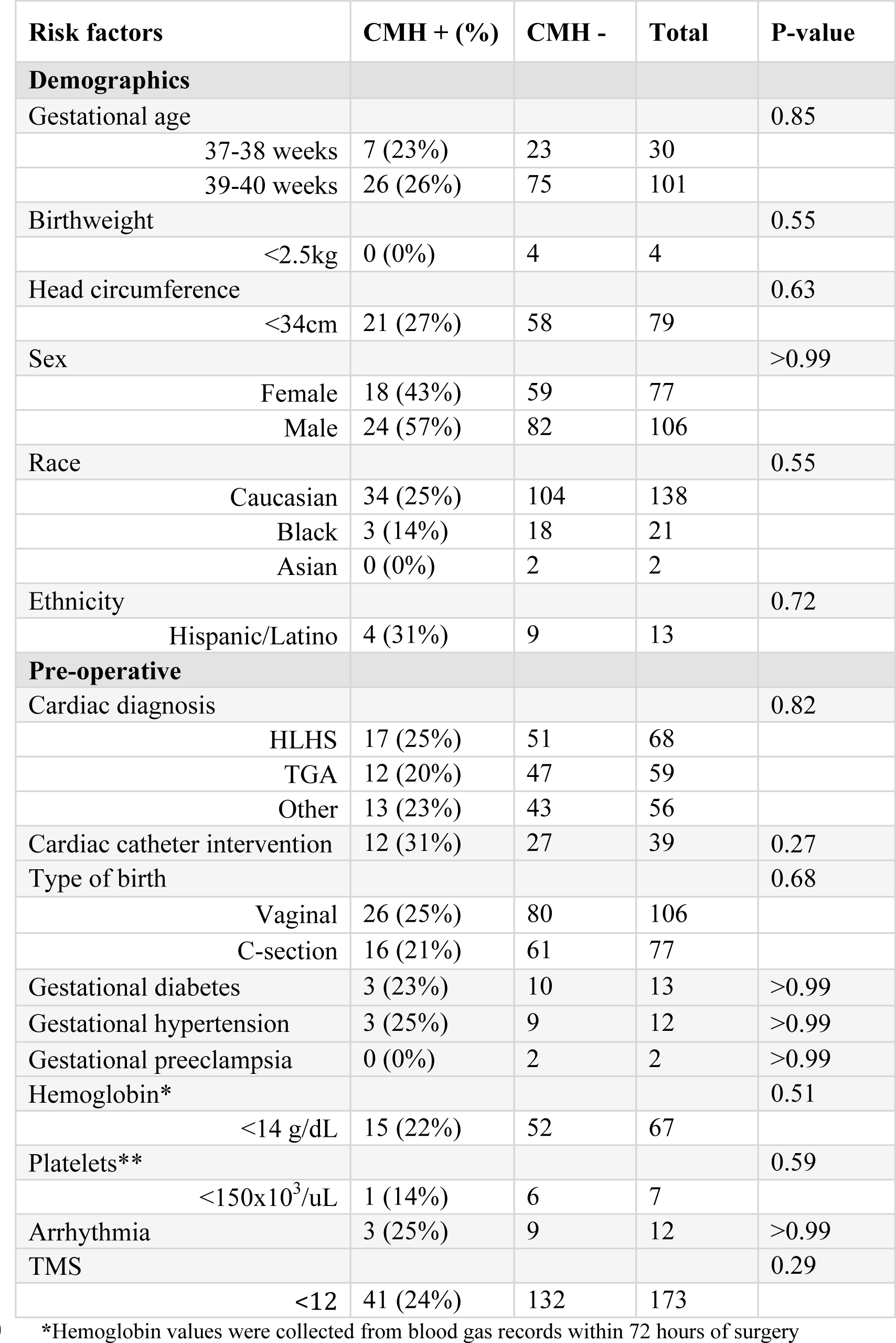
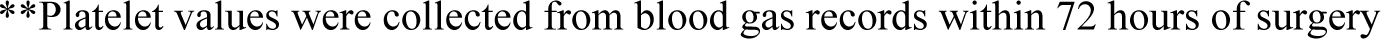
Association between candidate risk factors and cerebral microhemorrhage on preoperative magnetic resonance imaging (MRI). Fisher’s exact test was performed for categorical risk factors, whereas Wilcoxon rank sum test was performed for continuous risk factors. Cardiac catheter interventions included balloon atrial septostomy (BAS) and atrial stents. TMS, total maturation score.

| **Risk factors** | **CMH + (%)** | **CMH -** | **Total** | **P-value** |
| --- | --- | --- | --- | --- |
| **Demographics** |  |  |  |  |
| Gestational age |  |  |  | 0.85 |
| 37-38 weeks | 7 (23%) | 23 | 30 |  |
| 39-40 weeks | 26 (26%) | 75 | 101 |  |
| Birthweight |  |  |  | 0.55 |
| <2.5kg | 0 (0%) | 4 | 4 |  |
| Head circumference |  |  |  | 0.63 |
| <34cm | 21 (27%) | 58 | 79 |  |
| Sex |  |  |  | >0.99 |
| Female | 18 (43%) | 59 | 77 |  |
| Male | 24 (57%) | 82 | 106 |  |
| Race |  |  |  | 0.55 |
| Caucasian | 34 (25%) | 104 | 138 |  |
| Black | 3 (14%) | 18 | 21 |  |
| Asian | 0 (0%) | 2 | 2 |  |
| Ethnicity |  |  |  | 0.72 |
| Hispanic/Latino | 4 (31%) | 9 | 13 |  |
| **Pre-operative** |  |  |  |  |
| Cardiac diagnosis |  |  |  | 0.82 |
| HLHS | 17 (25%) | 51 | 68 |  |
| TGA | 12 (20%) | 47 | 59 |  |
| Other | 13 (23%) | 43 | 56 |  |
| Cardiac catheter intervention | 12 (31%) | 27 | 39 | 0.27 |
| Type of birth |  |  |  | 0.68 |
| Vaginal | 26 (25%) | 80 | 106 |  |
| C-section | 16 (21%) | 61 | 77 |  |
| Gestational diabetes | 3 (23%) | 10 | 13 | >0.99 |
| Gestational hypertension | 3 (25%) | 9 | 12 | >0.99 |
| Gestational preeclampsia | 0 (0%) | 2 | 2 | >0.99 |
| Hemoglobin* |  |  |  | 0.51 |
| <14 g/dL | 15 (22%) | 52 | 67 |  |
| Platelets** |  |  |  | 0.59 |
| <150x10<sup>3</sup>/uL | 1 (14%) | 6 | 7 |  |
| Arrhythmia | 3 (25%) | 9 | 12 | >0.99 |
| TMS |  |  |  | 0.29 |
| <12 | 41 (24%) | 132 | 173 |  |

\*Hemoglobin values were collected from blood gas records within 72 hours of surgery
\*\*Platelet values were collected from blood gas records within 72 hours of surgery

### Risk Factors for Postoperative Microhemorrhages

The median duration of CPB was 49 minutes [IQR= 39-74 min]. Longer duration of CPB was the risk factor most strongly associated with new or increased number of postoperative cerebral microhemorrhages (p<0.0001) (Figure 4a). Specifically, patients with postoperative cerebral microhemorrhages experienced a median CPB duration of 51 minutes [IQR= 40-84 min], compared to non-CMH patients who had a median CPB duration of 40 minutes [IQR= 30-44 min]. Additionally, need for postoperative ECMO support (p = 0.0004), presence of postoperative seizure(s) (p = 0.02), and low birth weight (defined as <2500g) (p = 0.03) were associated with worsened severity of postoperative cerebral microhemorrhage (Table 3). Age-at-surgery (p = 0.3), duration of DHCA (p = 0.3), lowest brain temperature (p = 0.98), maximum ACT (p = 0.2), intra-operative arrhythmia (p = 0.3), and postoperative cardiac arrest (p = 0.7) were not associated with change in number of cerebral microhemorrhage.

**Figure 4.**
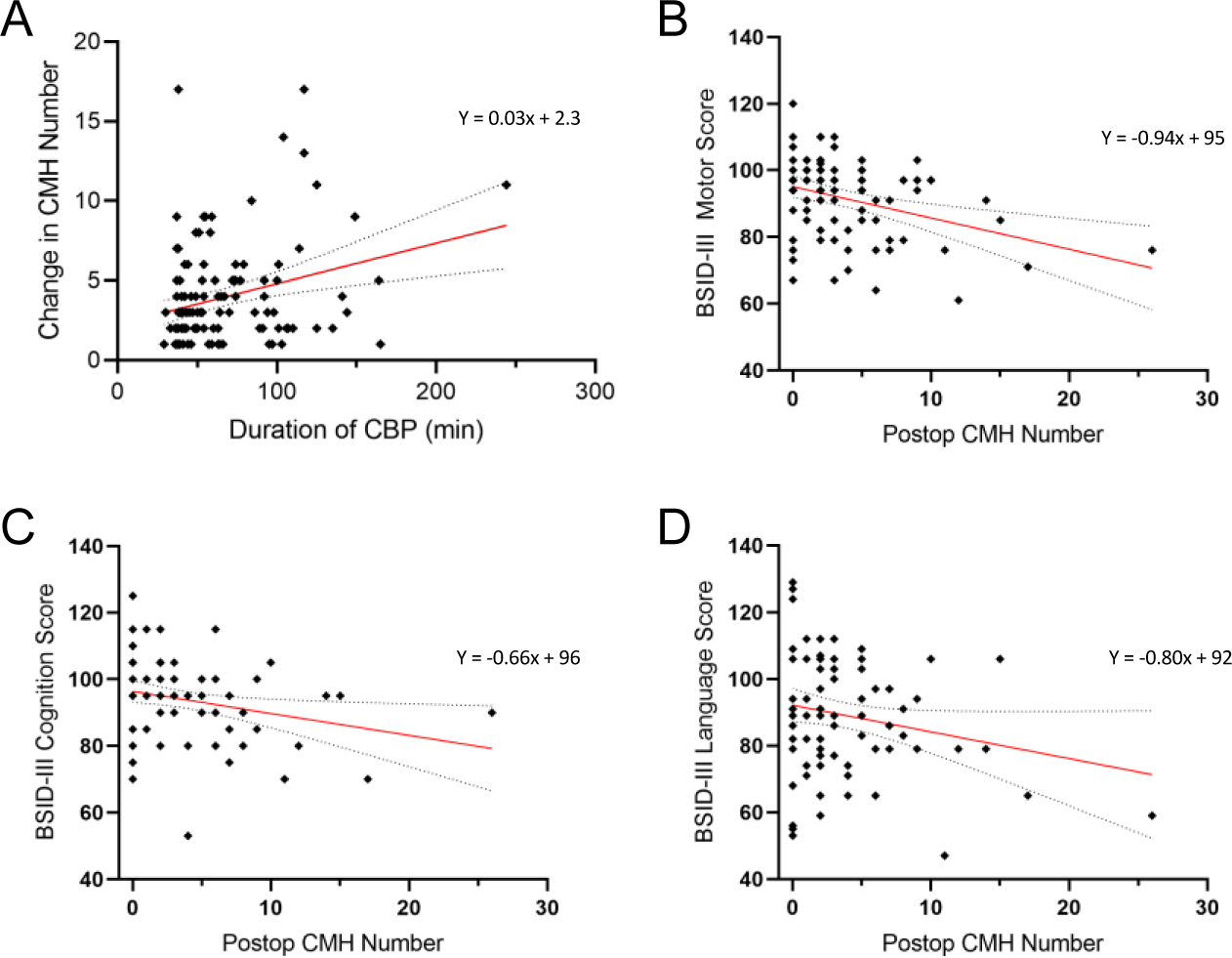
(a) Increased duration of cardiopulmonary bypass (CPB) is a significant risk factor for worsened cerebral microhemorrhage severity (p<0.0001). Cerebral microhemorrhage severity is defined as an increased number of CMH. **(b) The higher total number of cerebral microhemorrhages on postoperative scans is associated with lower composite motor scores at 18 months old (p=0.012) (c-d) Higher total number of cerebral microhemorrhages on postoperative scans is not associated with lower composite cognition (p=0.12) or language scores (p=0.06).** CMH indicates cerebral microhemorrhage; BSID-III indicates neurodevelopmental testing using Bayley-III Scales of Infant and Toddler Development at 18 months; postop, postoperative.

**Table 3.** Association between candidate risk factors and new or worsened cerebral microhemorrhage on postoperative magnetic resonance imaging (MRI). Linear regression was performed for both the univariate analysis and multivariate model. The multivariate model included birth weight and the significantly associated variables from univariate analyses.

| Risk factors | Beta (Std. Error) | P-value |
| --- | --- | --- |
|  | **Multivariate** |  |
| **Demographics** |  |  |
| Birthweight | -1.04 (0.479) | 0.03 |
| **Intra-operative** |  |  |
| Duration of bypass | 0.03 (0.007) | <0.0001 |
| **Postoperative** |  |  |
| ECMO support | 5.23 (1.44) | 0.0004 |
| Seizure | 1.94 (0.817) | 0.02 |

### Neurodevelopmental Outcomes

A subset of 82 patients with postoperative MRI underwent neurodevelopmental assessment using the Bayley-III Scales of Infants and Toddler Development at 18 months of age (Table 4). Neurodevelopmental testing revealed a wide range of composite scores for cognitive (53 - 125), motor (61 - 120), and language (47 - 129) function. The mean composite scores for cognitive, motor, and language function were 94 [SD± 12], 92 [SD± 12], and 89 [SD±16], respectively. While these scores indicate mid-average function for the cohort as a whole, 48% (n=39) of children in our cohort demonstrated below average neurodevelopmental function in at least one domain, defined as a Bayley’s score falling below 1SD of the mean. Additionally, 17% (n=14) showed pronounced impairment, falling below 2SD of the mean in at least one domain. Specifically, 5% (n=4) had moderate to severe cognitive impairment, 13% (n=11) had moderate to severe language impairment, and 5% (n=4) had moderate to severe motor impairment. Four patients had severe impairment in more than one domain, and one patient fell below 2SD of the mean in all three domains. Severity of cerebral microhemorrhage on postoperative MRI was associated with lower composite scores on the motor Bayley-III scales in both univariate (p=0.001) and multivariate (p=0.012) analysis (Figure 4b). The simple linear regression analysis suggested that with every one-point increase in CMH number, the Bayley-III motor score was predicted to decrease on average by 0.94 point (0.95% CI: 0.39-1.50). Lower composite cognitive scales were associated with cerebral microhemorrhage number on univariate (p=0.023) but not multivariate analysis (p = 0.118); use of postoperative ECMO was associated with lower composite cognitive score in multivariate analysis (p=0.007). The simple linear regression analysis suggested a 0.66 point decrease (95% CI: 0.09-1.22) in average Bayley-III cognitive score for every one-point increase in CMH number (Figure 4c).

**Table 4.** Association between candidate risk factors in patients who received postoperative magnetic resonance imaging (MRI) and composite cognition, language, and motor neurodevelopmental (ND) outcomes scores on Bayley-III Scales of Infant and Toddler Development at 18 months. Linear regression was performed for both univariate and then multivariate analyses. MultiV; multivariate. Postop; postoperative. GA; gestational age. CPB, cardiopulmonary bypass; ECMO, extracorporeal membrane oxygenation. WMI; white matter injury. QPS; quartered point score. TMS; total maturation score.

| Risk Factor | NEURODEVELOPMENTAL OUTCOME |  |  |  |  |  |
| --- | --- | --- | --- | --- | --- | --- |
|  | Cognition |  | Language |  | Motor |  |
|  | Univariate (p-value) | MultiV (p-value) | Univariate (p-value) | MultiV (p-value) | Univariate (p-value) | MultiV (p-value) |
| Postop CMH | 0.02 | 0.12 | 0.06 |  | 0.001 | 0.012 |
| Sex | 0.07 |  | 0.01 |  | 0.01 | 0.03 |
| GA | 0.46 |  | 0.45 |  | 0.81 |  |
| CPB duration | 0.49 |  | 0.20 |  | 0.59 |  |
| Postop ECMO | 0.002 | 0.007 | 0.007 | 0.02 | 0.07 |  |
| Seizure | 0.42 |  | 0.28 |  | 0.03 | 0.1 |
| WMI volume | 0.68 |  | 0.83 |  | 0.91 |  |
| QPS score | 0.35 |  | 0.86 |  | 0.50 |  |
| TMS | 0.99 |  | 0.42 |  | 0.59 |  |

Cerebral microhemorrhage was not associated with language function scores (p = 0.064), weight (p = 0.352), length (p = 0.712), or head-circumference (p = 0.185) in univariate analysis; these variables were not included in multivariate analyses. The slope was not significant in simple linear regression where the regression equation was y=-0.80x +92. Male sex was associated with a lower motor composite score on multivariate analysis (p = 0.027). In this cohort using simple linear regression analysis, WMI (total WMI volume and QPS score, respectively) was not associated with lower composite scores on cognition (p = 0.680; p = 0.352), motor (p = 0.906; p = 0.497), or language (p = 0.827; p = 0.857). Similarly, total maturation score (TMS) was not associated with lower composite scores on cognition (p = 0.989), motor (p = 0.595), or language (p = 0.421) in simple linear regression.

## Discussion

A major finding of our study is that CMH are not merely incidental findings but have clinical relevance as a prognostic imaging biomarker. The presence and severity of postoperative cerebral microhemorrhage are associated with impaired neurodevelopmental outcomes on the motor tests of the Bayley-III Scales of Infant and Toddler Development at 18 months of age. Specifically, for every one-point increase in CMH number, the Bayley-III motor score was predicted to decrease on average by 0.94 point (95% CI: 0.39-1.50) while the average Bayley-III cognitive score was predicted to decrease by 0.66 point (95% CI: 0.09-1.22). This finding is consistent with a prior study in this patient population that found that patients with cerebral microhemorrhage on MRI at one year of age had a 10-point lower score on the psychomotor development index (PDI) when compared to those without cerebral microhemorrhage.^22^ Our finding underscores the importance of reducing cerebral microhemorrhage burden.

### Risk factors for Preoperative CMH

The high prevalence of preoperative cerebral microhemorrhage in our cohort indicates that antenatal or preoperative factors contribute to risk for cerebral microhemorrhage. Although none of the antenatal and preoperative risk factors analyzed in this study was associated with the presence of preoperative cerebral microhemorrhage, we note that many of these analyses were underpowered to detect potentially meaningful differences. While previous studies have reported focal brain injury on preoperative MRI in infants with CHD who underwent the BAS procedure,^8,23^ catheter interventions on the atrial septum were not a predictor for CMH in our cohort. Stroke as a consequence of BAS was not seen in this cohort.

### Risk Factors for Postoperative CMH

We found longer duration of CPB to be the strongest predictor of an increased number of cerebral microhemorrhages on postoperative MRI. With CPB duration > 70 minutes, the median number of cerebral microhemorrhage was more than 2-fold higher compared to duration under 70 minutes. This finding is consistent with prior studies and may be the result of prolonged blood exposure to synthetic surfaces resulting in inflammation or embolic debris from the bypass machine.^24,25^ Moreover, the gross movement of brain tissue caused either by cerebral edema resulting from CPB or cerebral dehydration resulting from Lasix use may contribute to axonal stretching and the rupture of local vascular supply to affected brain tissue. This mechanism of injury is similar to that inflicted by trauma in diffuse axonal injury.^17,26^ Our analysis also showed that intra-operative factors including nadir temperature, duration of DHCA, and maximum activated clotting time (ACT) were not risk factors for cerebral microhemorrhages. We had hypothesized that deep hypothermia which can lead to brain shrinkage or water crystallization would be a risk factor, but our results did not support this.

In addition to the duration of CPB, the use of postoperative ECMO was also associated with increased postoperative CMH burden. Both CPB and ECMO are forms of extracorporeal support that likely share similar mechanisms of cerebral injury. As previously mentioned, systemic inflammation is triggered by prolonged patient contact with non-physiologic circuit components (i.e., pump and oxygenator) causing disruption to blood brain barrier integrity, and contributing to microvascular fragility.^27,28^ Infants with CHD have known structural brain immaturity^7,15^, and therefore may be more vulnerable to developing cerebral microhemorrhages with the use of extracorporeal support, as such interventions commonly impair cerebral autoregulation and cause hemodynamic instability that may further precipitate small vessel rupture.^27,29^ The use of anticoagulants with extracorporeal support may further contribute to cerebral microhemorrhage risk although we did not detect an association between intraoperative activated clotting time (ACT) and post-operative microhemorrhages in our cohort.^28^ Importantly, while these phenomena may explain the observed increase in postoperative microhemorrhages, they do not explain their presence preoperatively.

We report postoperative seizure and low birth weight as additional risk factors for postoperative cerebral microhemorrhage, which have not been previously reported to the best of our knowledge. It is not possible to determine whether seizure is a consequence or cause of cerebral microhemorrhage burden. However, as the prevalence of CMH was common (85%) and the prevalence of seizures relatively uncommon (9%), we believe them to be a consequence of CMH. Furthermore, prior research has shown that longer duration of CPB in infants undergoing CHD repair is a risk factor for postoperative seizure.^30,31^ Thus, both cerebral microhemorrhage and seizures may be sequelae due to a common underlying risk factor, though the presence of seizures may amplify the severity of the CMH. Seizures after neonatal cardiac surgery have been associated with worse long-term neurodevelopmental outcomes.^30,31^ Lower birth weight has previously been associated with higher mortality rates, poorer growth trajectories, and greater neurodevelopmental impairment.^19^ Future research investigating cerebral microhemorrhage in this population should evaluate the relationship between cerebral microhemorrhage, seizure, and lower birth weight.

### Pathophysiology

Cerebral microhemorrhage are most commonly reported in adult and elderly populations with several proposed underlying pathophysiologies. For example, diffuse axonal injury caused by rapid acceleration and deceleration of the brain in traumatic brain injury leads to mechanical stretching of axons.^26^ This stretching can trigger mitochondrial dysfunction, oxidative stress, and increased activity of metalloproteinases contributing to damage and dysfunction of the local microvasculature possibly giving rise to cerebral microhemorrhages.^26^ In contrast, cerebral microhemorrhages have also been widely detected in Alzheimer’s disease (AD) where they are thought to arise from inflammatory processes triggered by the accumulation of amyloid beta, tau, and cellular debris that characterize this disease.^32^ This proposed mechanism is supported by a previous study in AD patients that found an association between cerebral microhemorrhage and amyloid beta and tau load.^33^

We propose that a potential link between the mechanisms that give rise to cerebral microhemorrhages in adult and infant populations may be disruption to neurofluid balance via the glymphatic circulation. The glymphatic system is a waste clearance pathway within the brain that involves the exchange of cerebral spinal fluid (CSF) and interstitial fluid (ISF) to facilitate the removal of metabolic waste that accumulates in the brain during awake periods.^34^ Proteins and other metabolic waste products accumulate in the brain during waking hours due to the blood brain barrier which prevents their movement into systemic circulation and subsequent elimination from the body. Therefore, sleep is critical as a temporal regulator of the glymphatic system, facilitating fluid transport and promoting waste clearance.^34^ Disruption to metabolic waste removal via the glymphatic system has been implicated in Alzheimer’s disease (AD) and other neurodegenerative processes, where impaired glymphatic flow is thought to exacerbate the protein aggregation that is characteristic of these pathologies.^34,35^ Both CPB and DHCA may act to impede or disrupt normal neurofluid flow and metabolic waste clearance.

While cerebral microhemorrhage in the context of CHD is thought to be the result of debris and/ or inflammation from the CMB/ECMO circuit, this does not explain our observations of preoperative CMH. We propose that disruption to the glymphatic system and neurofluid clearance may contribute to CMH generation in this period. Glymphatic flow is driven by arterial pulsations generated by cardiac activity^34^ and therefore low cardiac ejection fraction, as seen in CHD infants, contributes to poor glymphatic flow and compromised metabolic waste clearance. Such disruptions to neurofluid circulation may leave the vasculature susceptible to oxidative stress and inflammatory processes that give rise to CMH via an AD-like mechanism. Alternatively, impaired neurofluid circulation may contribute to local swelling and dehydration of the brain that may give rise to CMH via a TBI-like mechanism.

Cardiovascular impairment for infants with CHD begins in utero and contributes to brain immaturity at birth with a delay of approximately 1 month of development.^7^ Such immaturity may be the result of placental insufficiency and a mismatch in oxygenation due to placental thrombosis or insufficient branching.^36,37^ This is supported by the recent finding that increased placental weight is protective against WMI.^11^ The glymphatic system may provide a link connecting cardiovascular risk factors to cerebrovascular health in this critical time period when there is enhanced fragility of small cerebral vessels which make them more vulnerable to developing pre-operative CMH. Understanding the role of the glymphatic system in cerebral microhemorrhage may provide insights into its underlying pathophysiology and provide potential therapeutic strategies to mitigate the clinical burden of these lesions. However, further research is needed to fully elucidate the role of the glymphatic system in cerebral microhemorrhage accrual in this population.

### Limitations

There are several limitations to this study. Although patients were prospectively enrolled, we were limited by the data available from this cohort in our retrospective analysis. Specifically, not all patients received both pre- and postoperative MRI. Only a subset of patients in our cohort (n = 82) returned for neurodevelopmental assessment, which could contribute to selection bias. Furthermore, there may be a contribution of genetic syndrome that is separately affecting neurodevelopmental outcomes, which could not be analyzed in this study due to the low numbers of infants with a known genetic diagnosis in our cohort.

We were also limited in our selection of risk factors to demographic factors and medical management factors that were relatively easy to abstract from the clinical record. Neonatal cardiac critical care is highly variable, and we were not able to capture all medications that were used (e.g., anticoagulants, pressors, or diuretics) or every clinical event (e.g., transient hypoxemia or apnea). There are likely many factors that contribute to cerebral microhemorrhage risk that are difficult to quantify. Further, at the time of patient recruitment, the preferred surgical strategy for aortic arch repair involved the use of DHCA, and it is unknown whether antegrade cerebral perfusion strategies are worse (longer CPB times) or protective for CMH.

Finally, our analysis of preoperative risk factors for cerebral microhemorrhage was underpowered to detect potentially meaningful associations. Future multi-center retrospective and prospective studies are needed to increase the sample size and help in better characterizing the risk of cerebral microhemorrhage in this population.

### Conclusions

Our data shows that cerebral microhemorrhage is a clinically significant problem in infants with severe CHD. We report both a high postoperative prevalence of cerebral microhemorrhage and an association between cerebral microhemorrhage on postoperative MRI and impaired neurodevelopmental outcomes at 18 months of age. Longer duration of CPB was the most significant risk factor associated with new or worsened postoperative cerebral microhemorrhage. Surgical factors, however, are not the only cause of cerebral microhemorrhage given the substantial preoperative prevalence observed. Further research is needed to better understand the mechanism of injury and to elucidate preoperative and postoperative cerebral microhemorrhage risk. The design of neuroprotection strategies to mitigate risk factors for cerebral microhemorrhage may improve neurodevelopmental outcomes in this vulnerable population.

## Data Availability

The data that supports the findings of this study are available upon reasonable request from the corresponding author.

## Acknowledgements

We extend our heartfelt appreciation to all those who contributed to this research including the Cardiac Kids Developmental Follow-up Program at CHOP. We are deeply grateful to the patients and their families who participated in this study and without whom this work would not be possible.

